# Seasonality and Progression of COVID-19 among Countries With or Without Lock-downs

**DOI:** 10.1101/2020.12.06.20244780

**Authors:** Jose-Luis Sagripanti

**Affiliations:** US Department of Defense Ringgold standard institution-US Army, US Army Combat Capabilities Development Command. Aberdeen Proving Ground, Maryland, United States

**Keywords:** SARS-CoV-2, COVID-19, coronaviruses, influenza, quarantine, stay-in-house, lock-down, mortality, epidemic, pandemic, virus inactivation, solar radiation, seasonal progression

## Abstract

Early predictions by computer simulation of 7 billion infections and 40 million deaths by COVID-19 during 2020 alone if lock-downs and other confining measures were not enforced may have justified restrictive policies mandated by governments of 165 countries. A main objective of the present study was to determine differences between the infection and death rate in countries that established early, nation-wide curfews, state-at-home orders, or lock-downs versus countries that did not mandate lock-downs to deal with the COVID-19 crisis.

The analyzed epidemiological data indicates that lock-downs, and other confining measures had no effect on the chances of healthy individuals becoming infected with- or dying of SARS-CoV-2.

The highest incidence of COVID-19 infection progressed from countries in northern latitudes, where it was winter at the beginning of the pandemic, to countries in the southern hemisphere in July 21, 2020 were winter was starting. This trend reversed again during the last quarter of 2020. A considerable (4-fold) increase in COVID-19 infection rate is observed between fall and beginning of winter in countries in the southern hemisphere. This seasonal progression correlates with the variation in the germicidal solar flux received by these countries, suggesting that infectious virus in the environment plays a role in the evolution of COVID-19. In addition, hypotheses are presented that could explain the recurrent new spikes of COVID-19 as well as the mortality of SARS-Co V-2 observed in some developed countries higher than the mortality rate reported in several developing countries.

## INTRODUCTION

The world has witnessed many epidemics in the past (McNeil, 1977). Other coronaviruses like SARS and MERS produced pandemics that started in 2002 and 2012, respectively (Peiris *et. al*., 2003; Ramadan and Shaib, 2019); and coronaviruses cause 15-20 % of all upper respiratory infections in humans, even in the absence of epidemics (Holmes, 1990). Several other viruses, like those of relevance in biodefense (with mortality rates of 40% for Lassa virus and 53% to 92% for the Sudan and Zaire strains of Ebola virus, respectively [Jahrling, 1997]) cause higher mortality than SARS-Co V-2 (global mortality rate of COVID-19 based on number of infections averaged 3.1% (with deaths per million approaching 0.012% globally) as of 22 September, 2020 [Johns’s Hopkins, 2020]). Rather what has been unusual were predictions made by computer modeling of 7 billion infections and 40 million deaths during 2020 alone if quarantine, lock-downs and other highly restrictive measures were not enforced (Walker *et. al*., 2020). These predictions may have been instrumental (Sagripanti, 2020) in justifying 1168 quarantine and lock-down policies mandated by governments of 165 countries (Cheng, C. *et. al*., 2020) that confined indoors at-risk as well as healthy individuals, resulting in a political and social crisis without historical precedents. Lock-downs and other restrictive measures remained in place in many countries throughout the year in spite of reports indicating as early as July-August 2020 that full lock downs were not associated with statistical significant reductions in the number of critical cases or overall mortality (Chaudhry *et. al*. 2020) and that infection rates and mortality rates felt among countries with and without lock-downs without a significant pattern (Sagripanti 2020). In addition, there have been a number of studies published during the last quarter of 2020 indicating that lock-downs or variable degrees of stay-at-home requirements were not statistically linked to- nor significant predictors of mortality, deaths per million or case fatality rate (Leffler at.al. 2020; DeLarochelambert et al 2020; Wehenkel, 2020). Moreover, any benefits of lock-downs were questioned in Germany (Wieland, 2020), not apparent in the Rep. of South Africa (Broadbent *et.al*., 2020), and ruled out as responsible in any decrease of the effective reproductive rate in the UK, indicating there that key predictions by computer simulation should be considered artifacts (Kuhbandner and Homburg 2020). Several of these findings could be considered highly technical, their findings regarding lock-downs sometimes unclear, preliminary, or confounded by additional variables or by complex scoring or indexes. A main aim of the present study was to clearly and rather straightforwardly identify any difference in infection or mortality as a result of nation-wide lock-downs by comparing progression of COVID-19 in two sets of countries and of US states (with or without lock-downs) through most of the year 2020.

It is well known that there is direct transmission of infectious virions by inhalation of contaminated aerosols exhaled, coughed, or sneezed from infected persons. Most measures mandated during COVID-19, from relatively benign (like wearing face masks or social distancing) to highly restrictive like quarantine, curfews, stay-at-home orders or lock-downs, were intended to prevent person-to-person transmission of disease which has been a main component in models used to predict the progression of viral epidemics (Walker *et.al*., 2020).

Direct (person-to-person) transmission was shown to be important in transmission of SARS-Co V-2 between nearby individuals (Chan, J.F. *et al*., 2020). However, it is remarkable that the COVID-19 pandemic progressed at a sustained rate after 26 March, 2020, when 1.7 billion people worldwide were under some form of indoor confinement, figure which increased to 3.9 billion people by the first week of April, amounting to more than half of the world’s population in quarantine or in-house lock-downs (Kaplan *et.al*. 2020; Sandford, 2020). However, the COVID-19 pandemic progressed unquenched in spite of greatly hindered person-to-person transmission.

The amount of infectious virus present in the aerosolized droplets produced by symptomatic patients or non-symptomatic carriers is not well established for coronaviruses, but it has been reported that nasal secretions contain up to 10^7^ infectious influenza viral particles per ml (Couch, 1995), from which aerosolized droplets generated by coughing, sneezing, and talking can contain several hundred infectious virions each (Gustin *et.al*., 2013). These micro droplets can reach distances of 12.5 meters (over 40 feet; Reiling, 2000). Coronaviruses have been reported to persist on contaminated surfaces with risk of disease transmission for up 9 days (Kramer *et.al*., 2006; Kampf *et.al*., 2020). SARS-CoV-2 persisted viable from 3 hours to 3 days depending on the type of surface on which it was deposited (vanDoremalen *et.al*., 2020). Influenza virus was readily re-aerosolized by sweeping floors without much loss in infectivity (Loosli *et.al*., 1943) and it should be assumed that SARS-CoV-2 will be reaerosolized in a similar manner. These findings indicate that SARS-CoV-2 should be able to persist for long periods in contaminated environments with continued risk of infection and progression of COVID-19.

Three main physical factors generally considered with a potential effect on virus persistence outdoors, include temperature, humidity, and the contribution of the germicidal (UVB) component in sunlight radiation. Laboratory experiments have demonstrated (particularly when virus infectivity was corrected by aerosol losses and natural decay) a rather limited effect of changes in relative humidity and ambient temperature on environmental virus survival and disease transmission (Hemmes *et.al*., 1960; Schaffer *et.al*., 1976; Kormuth *et.al*., 2018). Epidemiological studies on influenza concluded that the mortality increase in winter was largely independent of ambient temperature and humidity (Tiller *et.al*., 1983; Reichert *et.al*., 2004). Colder average monthly temperature was not associated with higher levels of COVID-19 mortality when accounting for other independent variables (Loeffler *et.al*., 2020).

Ultraviolet radiation in sunlight is the primary virucidal agent in the environment (Giese, 1976; Lytle and Sagripanti, 2005; Coohill and Sagripanti, 2009) and the relevance of sunlight in viral inactivation is documented (Mims, 2005; Sagripanti *et.al*., 2013). In contrast, smallpox, Ebola virus, Lassa virus, and other viruses of relevance in biodefense persisted in darkness for many days on contaminated surfaces (Downie and Dumbell, 1947; Sagripanti *et.al*., 2010). Modeling of viruses suspended in the atmosphere indicates that the diffuse (scatter) component of sunlight may still have approximately 50% of the virucidal efficacy exerted by direct solar radiation (Ben-David and Sagripanti, 2010; Ben-David and Sagripanti, 2013); demonstrating that viral inactivation by sunlight continues outdoors (albeit at half the rate or less) even in the shade or in polluted air or partially cloudy days. The UV sensitivity of coronaviruses, in general, and of SARS-CoV-2 in particular (Sagripanti and Lytle, 2020), indicates that 90% or more of SARS-CoV-2 virus should be inactivated after being exposed for 11-34 minutes of midday sunlight during summer in most world cities. In contrast, the virus will persist infectious for a day or more in winter (December-March in the northern hemisphere), with continued risk of re-arosolization and transmission at the same locations. Considering that SARS-CoV-2 could be three-times more sensitive to UV than influenza A (Sagripanti and Lytle, 2020; Sagripanti and Lytle, 2007), it should be inferred that sunlight should have an effect on coronaviruses transmission at least similar to that previously established for the evolution of influenza epidemics (Tiller *et. al*., 1983; Reichert et. al., 2004; Mims, 2005).

An understanding of COVID-19 should account for a) the rapid progression of the pandemic in spite of highly hindered opportunity for person-to-person contagion, b) the role of environmental transmission by contaminated aerosols, surfaces and fomites, c) a potential seasonal inactivation of virus in the environment by germicidal sunlight, and d) should explain the new spikes or waves of infections recurring even during summer, when germicidal sunlight radiation is more potent. This article attempts addressing these topics.

## METHODS

The total number of infections and deaths attributed to COVID-19 need to be normalized for comparative purposes (to everything being equal, large populations should have larger number of cases). Therefore, infection rate (infections attributed to COVID-19 per million inhabitants) and infection mortality rate (deaths per infections x 100) from different countries were considered in this study. However, infection mortality rates are strongly affected by the number of infections, which in turn varies with the level of testing. Serological testing uncovered relatively high numbers of asymptomatic cases, thus decreasing the infection fatality rate (Ioannidis, 2020). Therefore, the population mortality rate (deaths per million inhabitants) was also included in this analysis. Among available sources, the epidemiological data for the COVID-19 pandemics from John’s Hopkins’ Center for Systems Science and Engineering (John’s Hopkins, 2020) was employed. The reported mortality attributed to COVID-19 is not “excess mortality” as usually recorded in epidemiology. Therefore, the mortality figures used here could be an overestimation if basal mortality (mortality occurring in absence of epidemics) would be discounted. Countries reporting a progressive reduction of total deaths (fewer number of deaths after higher numbers) as well as countries were the number of reported COVID-19 infections decreased suddenly from rather high to very low, were considered unreliable and disregarded in this study. A map, as well as a list of countries and territories that did not mandate lock-downs can be freely downloaded from the world-wide-web (Wikipedia, 2020a); and a summary of countries with- and without lock-downs was also published elsewhere (Wehenkel, 2020) The infection per million figures were added for countries that instituted lock-downs and for “unlocked” countries that did not in each continent. The null hypothesis (no difference between countries with and without lock-downs) was tested (two tailed test, p < 0.01) for each paired set of data according to well established statistical analysis (Diem,1965; Wikipedia, 2020b) using an online calculator (Social Statistics, 2020).

The list of U.S. states and cities that ordered residents to stay at home has been previously reviewed (Mervosh et.al., 2020). The dates in which lock downs and quarantines came into effect (before 22 March, 29 March, 5 April, or 12 April, 2020) in various U.S. States have been monitored (Nguyen, 2020). Infection rate (cases per million inhabitants) in U.S. states was calculated by dividing the reported number of cases attributed to coronavirus (Statista, 2020; Centers for Disease Control and Prevention, 2020) by the state population (in millions to one decimal point) using the 2019 Census estimates taken by the United States Census Bureau (Wikipedia, 2020c). Graphs showing progression of COVID-19 used the specific data sets reported for the countries shown (John’s Hopkins, 2020; BBC News, 2020; Swedish Public Health Agency, 2020).

## RESULTS

### Confining people indoors

It has been estimated by computer modeling that without quarantine and lock-downs, the impact of COVID-19 pandemic would have been worse (Flaxman *et.al*. 2020; Hsiang *et.al*. 2020). The hypothesis may sound reasonable but if true, then, there should be a statistical significant difference between the infection rate and death rate in countries that established quarantine and lock-downs versus countries that did not mandated lock-downs to deal with the COVID-19 crisis. Some countries implemented nation-wide strict quarantine and in-house lock-down measures, often enforced by police, that decreased the time individuals could spent outdoors (Cheng *et. al*., 2020). Most countries implemented regional, less stringent lock-down measures at varying degrees. A third group of countries did not mandate lock-downs (Wikipedia, 2020a), thus freely allowing healthy individuals to remain outdoors with potential exposure to sunlight. These “unlocked” countries have not enforced any strict stay-at-home measures but have rather implemented large-scale social distancing, face mask wearing measures and/or instituted quarantine mainly for travelers and infected patients (Wikipedia, 2020a).

The benefit, if any, of quarantine and stay-at-home orders should be evident by comparing epidemiological data from countries with and without lock-downs. There is freely available and daily updated epidemiological data (John’s Hopkins, 2020) for countries, whose governments did not confine healthy citizen indoors (Wikipedia, 2020a). An analysis of the effect of in-house lock-down of healthy individuals in those countries that instituted partial or regional curfews or lock-downs is beyond the scope of the present study and it may be possible to attain only after COVID-19 had ended; however a limited but illustrative comparison among countries that imposed early the most restrictive lock-down measures and those that did not mandated healthy citizens to remain at home can be seen in Table 1.

**Table 1 A.**
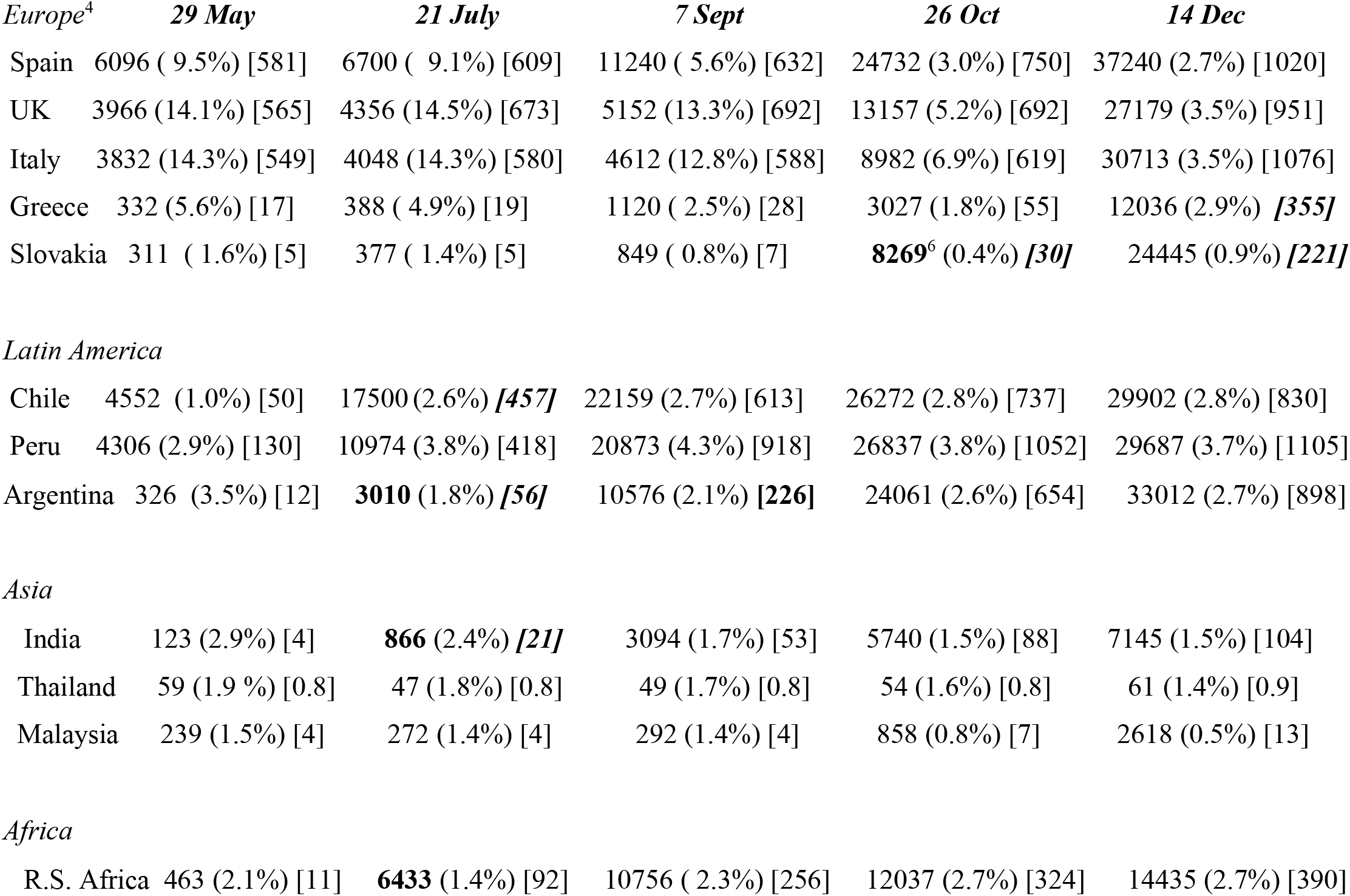
Countries^1^ that mandated Nation-Wide Lock-down. The Table shows infected per million^2^, mortality rate as (% of infected)^3^ and total deaths [per million inhabitants] attributed to COVID-19.

**Table 1 B.**
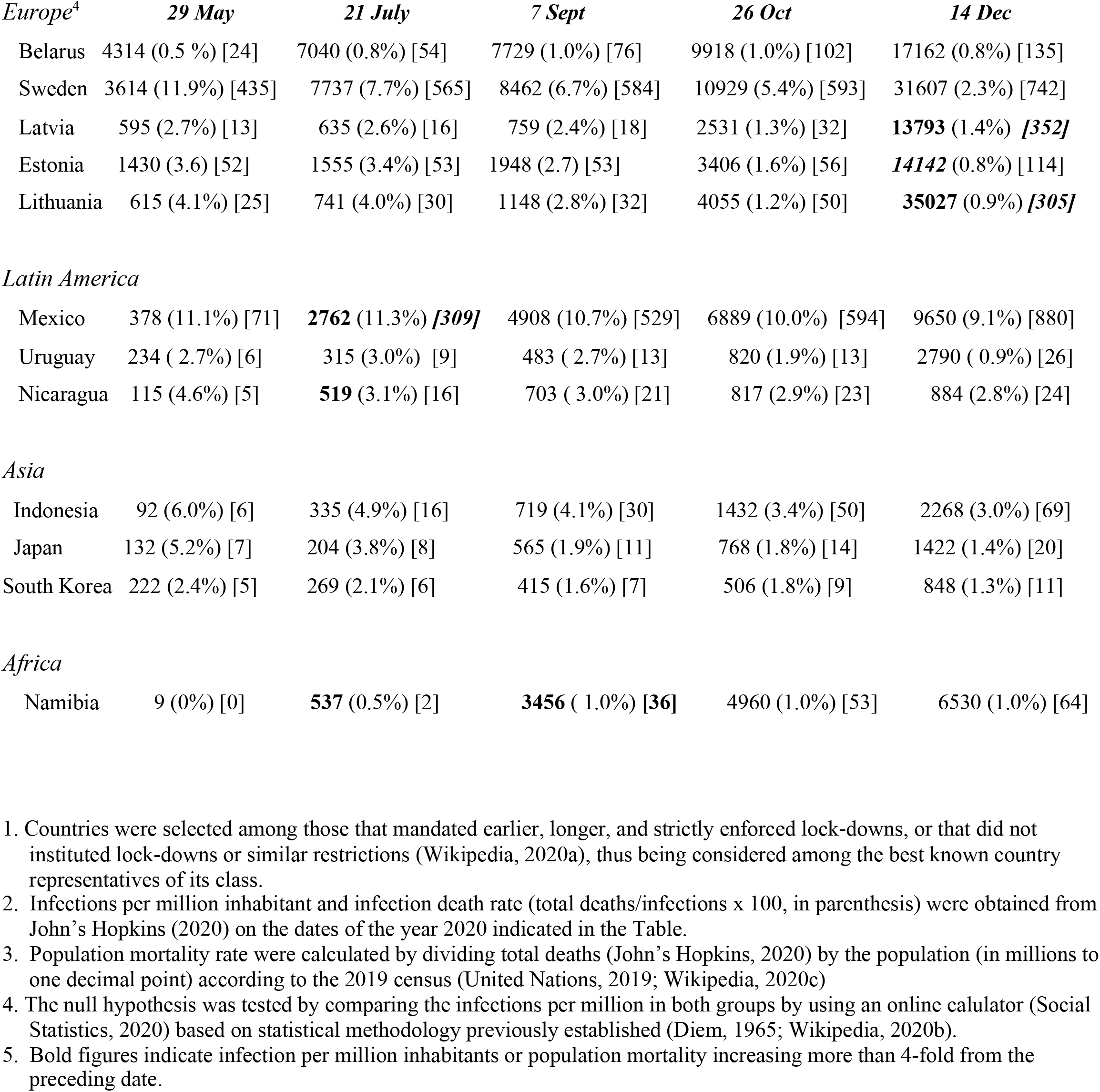
Countries Without Lock-down^1^.The Table shows infected per million, mortality rate as (% of infected)^2^ and total deaths [per million inhabitants]^3^ attributed to COVID-19.

Table 1 shows for each country the number of infections per million inhabitants and the national death rate as (percentage of infections) and as [deaths per million inhabitants] at various dates of the year 2020. The number of countries in the Table may be limited but they are among the best representatives of each group on each continent, have broad differences in the incidence of infection and mortality rates, and any significant difference resulting from quarantines and lock-down should be evident.

For example, among European countries showing in Table 1 the lowest infection incidence when summer was ending (7 September) Greece, who instituted most strict quarantine measures {1120/million, (2.5%), [28]} and Slovakia {849/million, (0.8%), [7]} which was among the first countries to mandate quarantine, actually have figures not too different to those in Latvia {759/million, (2.4%), [28]} or Lithuania {1148/million, 2.8, [32] which did not implement quarantine. Similarities can be seen in Table 1a and 1B also among countries with higher infection per million inhabitants in other continents.

The statistical analysis described in Methods demonstrated non-significant statistical difference in number of infections per million inhabitant between countries that instituted nation-wide stay-at-home orders versus countries that did not. Similar conclusions can be intuitively drawn by simply perusing the dispersion of data in Table 1 as well as the comprehensive epidemiological data base (John’s Hopkins, 2020).

The data reported for those states of the U.S. that instituted early lock-downs and states that did not confine the population indoors (Mervosh *et.al*., 2929; Nguyen, 2020) fails to show any statistically significant benefit of quarantine or lock-downs (Table 2). The data on 26 October, after the summer has ended, suggests a beneficial effect of stay-at-home orders (in average, around 40% less infections per million inhabitants among the analyzed US states that mandated lock-downs) but the difference is not statistically significant (p = 0.014714) within the constrains described in Methods (two-tailed T-test, p< 0.01). Similarly, there are no differences in infections between US states with- or without-lock-downs on 14 December, 2020 at the p<0.01 level (two tailed test, p-value is 0.116992).

**Table 2.**
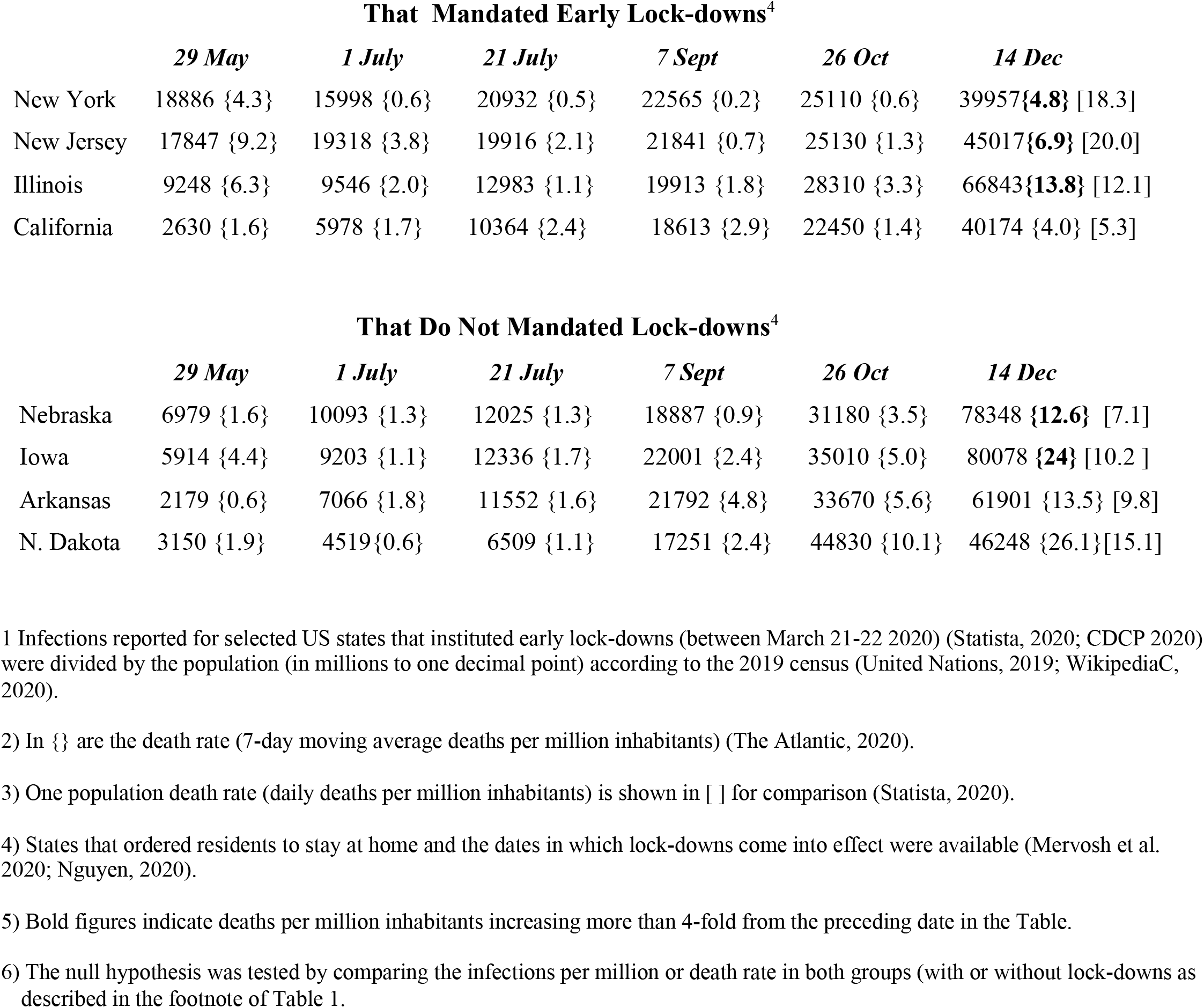
The table shows infection per million inhabitants^1^ and mortality rate^2,3^ attributed to COVID-19 in selected states of the U.S.A

### Progression of COVID-19

A progressive increase in infections per million inhabitants can be seen in Table 1 within the studied period for all countries (with or without lock-downs). The data in bold numbers presented in Table 1 indicates at least, a 4-fold increase in infections per million inhabitants between the subsequent dates shown. This considerable (4-fold) increase occurred in only one country (India) out of 12 represented countries in the Northern hemisphere between 29 May and 21 July, 2020. In contrast, all studied countries in South America and Africa, except one (Uruguay) reported at least a 4-fold increase between fall and beginning of winter in the southern hemisphere. These four fold increases in countries in the southern hemisphere occurred shortly after the date that these countries instituting lock-downs and other confining measures. The differences in relative increase of infections indicates a differential progression of COVID-19 between countries in northern and southern hemispheres following the respective seasonal shift from and into winter.

Moreover, the nearly two-fold increase (or more) in infection per million inhabitants recorded between 7 September and 26 October among all the European countries in Table 1 that instituted lock-downs correlates with the solar progression from summer into winter.

Four fold increases in mortality per million inhabitants (bold in Table 1) occurred during the fall (October-December) in Greece and Slovakia as well as in Latvia and Lithuania. Four fold increases in infections occurred also during the fall in the southern hemisphere (May-July) after Chile had imposed partial curfews and Peru and Argentina initiated stringent stay-at-home orders between 16-19 March 19th (Wikipedia, 202a; Larrrosa, 2020).

According to data reported on May 7, 2020 (John’s Hopkins, 2020), of the 30 countries with highest COVID-19 infections per million inhabitants, 28 were located north of the Tropic of Cancer (the two exceptions being equatorial Qatar and Mayotte at latitudes 25° N and −13° S, respectively).

The epidemiological data reported on July 21, 2020 from the same source shows that the composition of the 30 countries with highest incidence had changed. On July 21, of the 30 countries with highest infection per million inhabitants only 14 countries where still located in the northern hemisphere, 11 countries were equatorial (located within latitude +/− 26°) and 5 countries (Chile, Peru, Brazil, Bolivia and South Africa) were located in the southern hemisphere. These data indicates that the highest incidence of COVID-19 infection progressed from countries in northern latitudes, where it was winter at the beginning of the pandemic, to countries in the southern hemisphere were was winter in July 21. This seasonal progression correlates with the variation in the virucidal solar flux received by several of these countries previously reported (Sagripanti and Lytle, 2020). As of 7 September 2020, the location of the 30 countries with highest incidence of COVID-19 has dispersed north and south of the equator without an obvious geographical pattern. The majority (14 of the 30) of countries with highest infection per million inhabitant on 26 October, 2020 (John’s Hopkins, 2020) are located near the equator, consistent with solar transit to the equinox. On 14 December the composition of the 30 countries with highest number of infections has changed again, with only six countries in the southern hemisphere where summer is starting and 24 countries in the northern hemisphere where sunlight radiation has been decreasing due to incoming winter. Twenty eight of these 30 countries with highest infections numbers on 14 December mandated lock-downs or instituted stay-home orders (the two countries that did not ordered lock-downs among these being Mexico and Indonesia)

### New spikes of infections

An understanding of COVID-19 must include a plausible explanation for the new waves of infections that resulted in spikes in the epidemiological data. Figure 1 shows the number of active cases as a function of date according to national reports (John’s Hopkins, 2020) from representative European countries listed in Table 1. Countries in Fig 1A shows a bi-modal distribution with a first peak around mid-April 2020, and a valley either in mid-May, early May or mid-June in Spain, Greece and Slovakia, respectively while the number of active COVID-19 cases spike in the three countries after that date. Greece was among the earliest countries to mandate lock downs and Spain had among the most restrictive lock down measures in Europe. For example, Spain mandated lockdown early in 27 March, partially relaxed some restrictions on 13 April, about a month before a new wave of infections begun (BBC News, 2020). In contrast, the European countries presented in Fig. 1B (John’s Hopkins, 2020, Swedish Public Agency, 2020) that did not mandate lock-downs and allowed healthy people to remain outdoor (Wikipedia, 2020a) do not show a bimodal curve of COVID-19 like those in Fig 1A.

**Figure 1.**
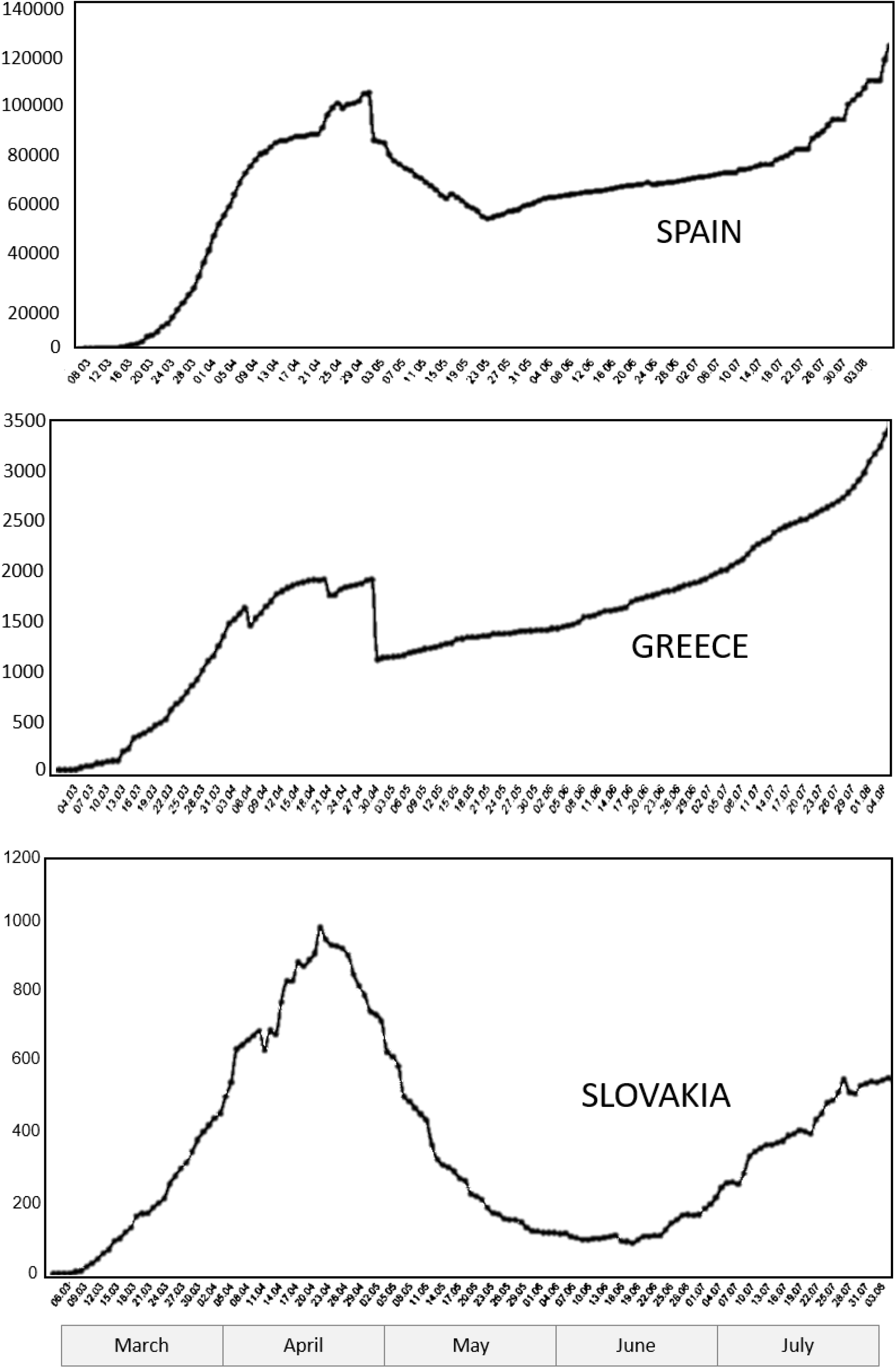
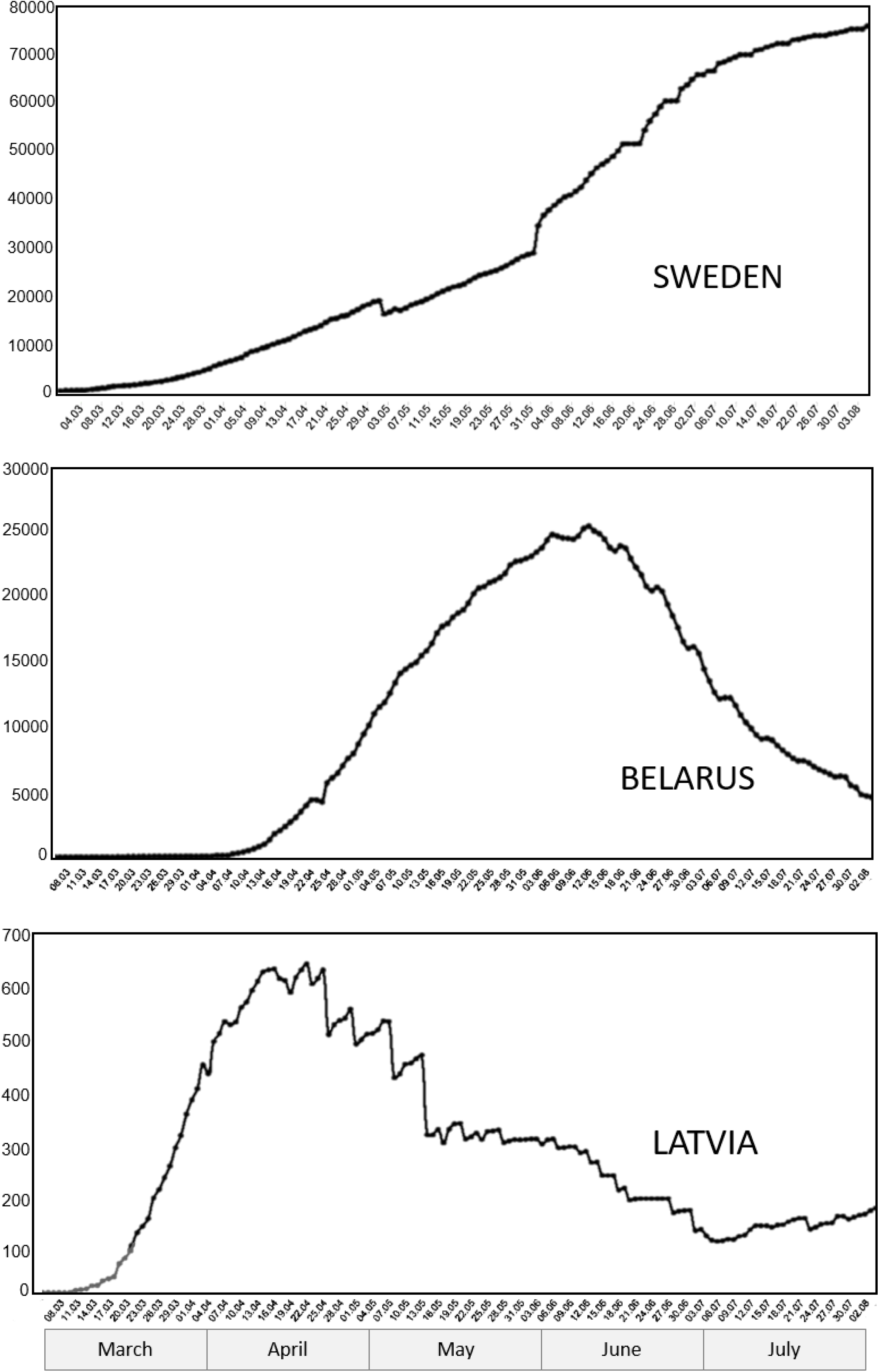
Number of active cases as a function of date from representative European countries listed in Table 1; (A) that mandated lock-downs and (B) that did not mandated lock-downs

Figures reported for at least 5 US states shown in Table 2 indicates that lock-downs did not prevent the spikes resulting in a four-fold increase in population mortality rate observed between 26 October and 14 December (in bold), when summer had ended and winter was approaching.

A sharp increase in number of infected and active cases occurred at the beginning of September 2020 in Spain, the middle of September in Greece, late September in France, UK, and Slovakia, and early October 2020 in Italy (John’s Hopkins, 2020) (data not shown), demonstrating that lock-downs failed to prevent progression of COVID-19, at least after summer in Europe was ending.

Not all European countries in Table 1A that mandated lock-downs show a bimodal curve within the period studied. Only Uruguay and Japan that did not mandate lock-downs show a bimodal progression for COVID-19. Several Asian countries and most countries in the southern hemisphere listed in Table 1 have reports of daily cases that vary rather wildly as a function of date, without the necessary accuracy to clearly determine accurately the shape of COVID-19 progression.

Interestingly, most of the US states that instituted stay-home orders did so from 19 March (a week to ten days before the new infections peak). Stay-home orders were partially lifted by many U.S. states in early-mid May (Nguyen, 2020) during the valley of daily infections (data not shown). The lifting or relaxing of stay-home orders seems to precede and correlate with COVID-19 expansion.

### Mortality rate

Mortality rates among countries in Table 1A with and 1B fail to show a significant differences between the two groups of countries, demonstrating that lock-downs and stay-home orders do not determine the mortality rate of COVID-19. However, the infection mortality rates for COVID-19 (deaths over infections x 100) shown in Table 1 vary dramatically, ranging from less than 1% to over 14%) and population mortality rates [deaths per million inhabitants] as late as 14 December range between 1 and 1100 among different countries (John’s Hopkins, 2020).

As of July 21 there were 17 countries with infection mortality rates at least 2-fold higher than the world average at that time (4.1%). From these countries, 9 were relatively small developing countries (4 of which are small islands) with health care difficult to assess. The remaining 8 countries with death rates more than 8.2% included France (15.9%), Belgium (15.3%), UK (14.5%), Italy (14.3 %), Hungary (13.7%), Netherlands (11.8%), Mexico (11.3%) and Spain (9.1%). All of these countries are located in the northern hemisphere where it was winter at the beginning of the pandemic. Only one of these, Mexico, did not mandate lock-downs of healthy individuals. Sweden that has been considered representative of “unlocked country” had a mortality rate of 7.2% (slightly inferior to the two-fold average of 8.2%). As of 7 September, 2020, Sweden’s infection per million and infection mortality rate remained considerably higher (8462, 6.7%) than its Scandinavian neighbors that instituted lock-downs (Norway 2098, 2.3% and Finland 1502, 4%). Although infection mortality rates have decreased in 26 October, Sweden mortality rate remains higher (5.1%) than its neighbors (1.5% and 2.3% for Norway and Finland, respectively). However, these differences could diminish if adjusted by the chances of contagion related to population density (in people per km^2^: 22 for Sweden versus 15 for both Norway and Finland) or to the crowding in their larger cities (with Stockholm nearly three times more populous than Oslo or Helsinki [World Population Reviews, 2020]). The population mortality rate (deaths per million inhabitants) of Sweden (Table 1B) has trailed behind countries like Spain, the UK and Italy that established early nation-wide lock-downs (Table 1A).

In contrast with considerable increases observed in infections per million inhabitants, infection mortality rate in each country was relatively constant between 29 May and 26 October, 2020 (Table 1). This observation suggests that the mortality resulting from COVID-19 is independent from seasonal sunlight and instead, large disparity of mortality for the same virus among different countries could correlate to variables yet to be elucidated. The infection mortality rate (as of 7 September, 2020) among every major developing country in South America (those countries shown in Table 1 plus Colombia 3.2%, Brazil 3.1%, Paraguay 1.9%, and Venezuela 0.8%) is considerably lower than the infection mortality rate in several developed countries in Europe. Exceptions of Latin American countries with relatively high mortality rates like Mexico and Ecuador (infection mortality rate 9%) are at- or north of-the equator. The population mortality rate of the three Latin American countries in Fig 1A that instituted nation-wide lock-downs (Chile, Peru, and Argentina) is similar to that of Spain, the UK, and Italy, and higher than in Uruguay and Nicaragua (Latin American countries in the southern hemisphere that did not mandate lock-downs) (Table 1B).

## DISCUSSION

Predicting 7 billion infections and 40 million deaths in 2020 alone (Walker *et.al*., 2020) suggests that current modeling of epidemics could be omitting a variable of importance in limiting the progression of viral pandemics (Ioannidis *et.al*. 2020; Saltelli, 2020; Sagripanti, 2020). Further estimating by computer modeling that millions of deaths were averted by lock-downs (Flaxman, *et.al*., 2020; Hsiang, *et.al*., 2020) strongly disagrees with the epidemiological data compiled by John’s Hopkins (2020) and with the global epidemiological data analyzed here. To justify the high cost of lock-downs, any positive effect should have clearly (and statistically significantly) surfaced above any potential co-founding variable (mask wearing, obesity, etc.). The fact that the rather simple and straightforward approach employed here (key data in two sets of countries followed through most of the year) failed to detect any benefit in COVID-19 infection or mortality in countries that instituted nation-wide lock-downs should be convincing evidence. Any limitations assigned to the accuracy of the data reported by different countries or US states should affect equally both data sets (with and without lock-downs), thus having little bearing on the conclusions being presented.

The lack of a positive effect of lock-downs presented here agrees with previous studies using virtual simulation as well as analysis of epidemiological data reporting that lock downs were both superfluous (did not prevent the explosive spread of COVID-19) and ineffective (did not slow down the death growth rate) (Chaudhry *et.al*., 2020, Sagripanti, 2020; Kuhbandner and Homburg, 2020; DeLarochelambert *et.al*., 2020; Wehenkel, 2020; Leffler et.al. 2020). These findings demonstrate by different and independent approaches that virtual simulation of epidemics should complement but never replace actual epidemiological data in policy making.

The presented data (Table 1) indicates that COVID-19 progressed differently in countries at northern latitudes as it was winter time and sun exposure was limited at the onset of the pandemic during December 2019-March 2020, than in countries in the southern latitudes where summer sunlight was abundant. The shift of highest infection rates from countries in the northern-towards countries in the southern-hemisphere reported in the present work correlates with seasonal variation in the flux of germicidal sunlight (Lytle and Sagripanti, 2005; Sagripanti and Lytle, 2007; Sagripanti and Lytle 2020). Together with the limited virucidal effect (within the ambient range) reported for temperature and humidity (Hemmes *et.al*., 1960; Schaffer *et.al*., 1976; Kormuth *et.al*., 2018; Tiller *et.al*., 1983; Reichert *et.al*., 2004, DeLarochelambert, 2020), the most plausible explanation for the observed north-south shift of infection is the role of virucidal sunlight in the progression of COVID-19 (Sagripanti and Lytle, 2020).

Although not every country in the world and state of the USA is discussed in the present study, the countries considered (24 countries analyzed in Table 1 plus 13 additional countries addressed in the text of Results, plus 8 US states) should be enough to detect significant differences resulting from quarantines and lock-downs if significant differences should exist. Less definitive should be considered a correlation between lock-downs and a bimodal progression of COVID-19. However, the curves shown for at least some European countries (Fig 1A) early in the pandemic suggests that mandating individuals to remain indoors may have altered the natural progression of COVID-19. Quarantine and other restrictive confining measures may have worsened the prospects of certain countries, particularly of those in the southern hemisphere that rushed to emulate the policies taken by countries in the northern hemisphere without considering the seasonal difference. Quarantine in the southern hemisphere was implemented in several countries during early 2020, summer time there, where the sun at full virucidal potential could have reduced the progression of the epidemic by naturally reducing the infectious viral load in the environment (Sagripanti and Lytle, 2020) broadcasted by the then relatively low number of cases in these countries (John’s Hopkins, 2020). Later (in July 2020) several of these southern hemispheric countries still implementing strict quarantines (see Table 1) entered winter with a considerable population of susceptible individuals that, when eventually released from quarantine, could fuel COVID-19. However, at this time, countries in the southern hemisphere did not have intense sunlight to rapidly inactivate the virus in contaminated environments (Sagripanti and Lytle, 2020) thus accounting for the relatively large increase in infections (bold figures in Table 1).

The data presented herein indicates at least three potential factors that could increase the number of COVID-19 infections. The most obvious is a seasonal component in the pandemic correlated to local solar flux. This component demonstrates that virus infectious in the environment plays a role in the pandemic since direct person-to-person transmission should be relatively independent of solar inactivation.

A second factor can be identified among some countries where an increase in infections (and mortality) occurred even after lock-downs and other stringent confining measures had been in place. This increase could be related to confining people indoors, at home or in nursing homes, thus increasing (or assuring) contagion among individuals under the same roof.

Lastly, spikes of COVID-19 can be detected after stay-home sanctions are lifted. Like wildfires feeding off dry timber and hurricanes hot moist air, epidemics need susceptible individuals to persist and progress. The end of quarantines could fuel COVID-19 spikes when a considerable population of susceptible individuals kept locked in their homes, (thus deprived of sunlight, lowered their levels of vitamin D, and weakened their immune competence by staying long periods indoor [Burren *et.al*., 2015; Geddes, 2020]) are eventually released, therefore increasing the chances for COVID-19 to flash up. This possibility seems supported by previous data indicating that lock-down in Argentina (among the longest and more strict in the world) only postponed infections by hindering mobility (Larrosa, 2020). Strict curfews and lock-downs did not prevent Argentina from increasing 4-fold its mortality rate twice (from May to September) as shown in Table 1A (in bold).

In contrast with considerable increases in infection per million inhabitants, relatively constant infection mortality rate in each country [between 29 May and 14 December, 2020 (Table 1)] suggest that the mortality resulting from COVID-19 infections is independent from seasonal sunlight and instead, mortality after infection should correlate with national health characteristics.

COVID-19 infection mortality rates in developing countries of south America (11 of the largest countries) was considerably lower than in several (at least 8) developed European countries (see Results). Although perhaps counter-intuitive, this finding agrees with a previous report correlating higher national Gross Domestic Product (GDP) with higher mortality rates (DeLarochelambert *et.al*., 2020). National differences in infection mortality rate (at times 4-fold or higher than the global average) produced by the same virus could result from a less efficient national health care system (due to lack of investment, training, or proper allocation of relevant resources). However, this notion seems at odds with the perceived differences in resources and health budget between European and South American countries. It has been suggested that an older and more sedentary population in countries with higher GDP could account for higher mortality rate than that in developing countries (DeLarochelambert *et.al*., 2020). Alternatively, it could be speculated that higher mortality rates after infection by the same virus in developed countries could be the result of a more susceptible population (due, for example, to consumption of highly processed food or exposure to artificial antigens and pollutants) than the population in developing countries (generally ingesting a diet of unprocessed food and living in more natural environments). This seem supported by the strong correlation previously reported between obesity and COVID-19 case load (Chaudhry et al, 2020; DeLarochelambert *et.al*., 2020). However, the reasons behind differential mortality rates between developed and developing countries needs extensive elucidation.

Considering the enormous number of infectious particles that can be broadcasted by an infected person (Couch, 1995), even during the incubation period before symptoms can be detected (when molecular tests are still insensitive), the unavoidable contamination of environments indoor as well as outdoors must be considered (particularly in modeling) as a viral reservoir throughout the epidemic. This reservoir should be encompassed by a wide variety of contaminated natural and man-made surfaces and fomites, where SARS-CoV-2 can be inactivated relatively rapidly by summer sunlight or can persist with risk of infection for long periods at higher latitudes during winter or indoors most of the year (Sagripanti and Lytle, 2020).

We can envision the progression of the epidemic as the result of the interaction and mutual balance of at least four main factors (some counteracting each other). The first involves the virulence and contagiousness of the germ, being slow but progressively attenuated by multiple passages through healthy immune-competent hosts. Secondly, this virulence is more or less countered by the higher or lesser capability of the human host in mounting an efficient immune-response. The human host sensitivity to infection fluctuating with the patient general health, nutrition, metabolic condition (including levels of active vitamin D), expression of relevant genes, and psychological outlook of each individual during the crisis (Burren *et.al*., 2015; Geddes, 2020). Third, throughout the epidemic, a considerable virus load is broadcasted by infected patients (even asymptomatic) into the environment, contaminating a variety of surfaces. Lastly, fourth, in the absence of sunlight (during winter in many temperate zones and indoors), the reservoir of virus in contaminated environments persist with risk of infectious for considerable periods of time. In contrast, when sunlight is abundant, viral inactivation proceeds rather quickly even in the shade (Lytle and Sagripanti, 2005; Sagripanti et.al., 2013; Ben-David and Sagripanti, 2010; Ben-David and Sagripanti, 2013; Sagripanti and Lytle, 2020).

Similar epidemiological data amongst “locked” and “unlocked” countries presented in this study demonstrate that measures confining populations indoors by lock-downs had no effect on the chances of healthy individuals becoming infected with SARS-CoV-2, or dying of COVID-19. Given the importance for pandemics certainly to come, albeit in an uncertain future, the inability of quarantines and lock-downs on controlling the progression of COVID-19 should not be discarded lightly, not at least without consulting the freely available epidemiological data (John’s Hopkins, 2020).

## CONCLUSION

The differential increase in infections per million inhabitants between countries in the northern versus the southern hemisphere indicates a seasonal component in the progression of COVID-19. This seasonal progression indicates that an environmental component plays a relevant role in the pandemic. Mortality rate for the same virus among different countries did not show a seasonal component and instead, it was relatively characteristic for each country. When lock-downs are eventually lifted, susceptible individuals exposed to virus persisting in the environment could fuel new spikes of infection. The data presented and arguments discussed here indicate that lock-downs and other restrictive measures confining at-risk as well as healthy individuals indoors, may have failed in preventing, or significantly reducing, the spread or mortality of COVID-19 as such policies intended.

## Data Availability

The epidemiological data employed in this study is freely available from the sources listed in References. Most reprint of the listed References can be downloaded from the world wide web. Those articles not freely available can be requested from the author.

## Acknowledgements

*The critical review of Dr. C.D.Lytle, the graphics support of Mrs. D.M. Hoffman, end the editorial review by Mrs. F. Zelachowski are highly appreciated*.

## Author contribution

*The single author (Dr. Jose-Luis Sagripanti) is sole responsible for the content of the article*.

## Data Availability Statement

*The epidemiological data employed in this study is freely available from the sources listed in References. Most reprint of the listed References can be downloaded from the world wide web. Those articles not freely available can be requested from the author*.

## Animal and Human Experiments

*No experiments utilizing animals or humans were undertaken in this study*

